# The Effects of Intermittent Vibrational Stimulation on Lower Limb Kinematics Two Months After Anterior Cruciate Ligament Reconstruction: A Randomized Controlled Trial

**DOI:** 10.1101/2024.05.13.24307234

**Authors:** Tomer Yona, Bezalel Peskin, Arielle Fischer

## Abstract

**Objective:** To investigate the immediate effects of intermittent vibrational stimulation on gait and stair ambulation among patients two months post anterior cruciate ligament reconstruction.

**Design:** Randomized, parallel, 2-group-randomized controlled trial.

**Setting:** Hospital setting.

**Participants:** There were 27 male and female participants aged 18-45, two months post-ACLR, and a convenience sample of 24 healthy controls.

**Interventions:** The participants were randomly assigned to two groups. One group received a device designed to apply intermittent vibrational stimulation above and below the knee; the other received a sham device.

**Main Outcome Measure(s):** The main outcomes were the knee sagittal kinematics during gait at three speeds and stair ascent and descent. The assessment was performed with and without one of the study devices.

**Results:** Compared to the sham device, intermittent vibrational stimulation significantly increased the minimum knee flexion angles while walking at normal, slow, and fast paces and stair ascent.

**Concblusion:** Intermittent vibrational stimulation affects the lower limb kinematics during rehabilitation for individuals post-ACLR. However, further research is necessary to confirm long-term benefits and establish optimal application parameters.

## Introduction

Anterior cruciate ligament (ACL) injuries are a prevalent concern, especially among active young people.^1^ Such injuries lead to extended changes in everyday life and sports activities. They are followed by a lengthy rehabilitation process, usually lasting at least nine months before returning to the activity levels before the injury.^2^ Despite rehabilitation efforts, many individuals still experience persistent ambulatory changes in everyday activities such as walking and ascending or descending stairs.^3–5^

The reported ambulatory alterations after ACL injury have clinical implications, indicating a need for exploring novel approaches to address these changes. Quadriceps weakness post-ACL injury first associated with ‘quadriceps avoidance gait’ by Berchuck et al. (1990)^6^ can present a barrier to rehabilitation. This type of gait can cause asymmetry and reduced walking speed, leading to changes in joint movement patterns, affecting joint mechanics and loading, and potentially contributing to osteoarthritis (OA).^7,8^ Increased risk of developing premature OA presents a long-term and unresolved risk following ACL injury and motivates to seek new approaches to mitigate this risk. The gait changes noted above, as well as the risk of developing OA, suggest the need for modifiable gait metrics associated with premature OA.

A new intervention (*KneeMo^®^*) uses intermittent vibrational (IVS) to apply to sensory nerves that signal pain as well as mechanical stimuli such as vibration. It uses the properties of the somatosensory system to gate pain and enhance function, as previously described in detail.^3,9,10^ However, the specific effects of IVS on the kinematics of the lower limb following ACLR, particularly during the early phase of rehabilitation, remain inadequately explored.

While walking has received considerable attention in ACL rehabilitation research, the biomechanics of stair ambulation remain relatively understudied.^4^ Yet, there is evidence that joint loading and knee flexion are altered during stair use after ACLR surgery, highlighting the need for early interventions.^11,10^

The importance and challenges of early-phase rehabilitation have been previously described and provide the motivation for evaluating new interventions. Specifically, Buckthorpe et al. (2020) have reported that mid-stage rehabilitation after ACLR is an important timeframe for interventions addressing altered gait patterns to prevent long-term limitations.^12^ Implementing gait retraining techniques during this phase can improve aberrant gait patterns. Therefore, investigating the effects of IVS at the two-month post-ACLR mark is particularly important to assess its efficacy as an early rehabilitative intervention. Moreover, employing a sham device as a control helps reduce potential placebo effects, where participants may anticipate benefits solely from receiving an intervention. By comparing the effects between the groups, we can more confidently attribute any observed improvements to the vibration device.

Traditionally, kinematic analysis is done using optoelectronic camera-based motion capture systems within the controlled environment of a movement laboratory. While accurate, it can potentially limit the ecological validity. Inertial measurement units (IMUs) present a promising alternative, enabling kinematics data collection outside the lab setting.

We aim to address this research gap by evaluating the influence of IVS on knee kinematics in individuals two months post-ACL reconstruction using IMUs outside the laboratory. Our primary objective is to assess the effects of IVS on the knee sagittal plane movements during overground walking and stair ascent and descent. Additionally, we aim to examine the sagittal plane movements of the hip and ankle during these movements. We hypothesized that individuals receiving a device applying IVS to the knee during ambulation would demonstrate increased knee flexion angles compared to those in the sham device group.

## Methods

### Study design

The study was a randomized controlled trial with a parallel 2-group before and after design and followed the Consolidated Standards of Reporting Trials (CONSORT) Statement.^13^

The institutional review board of Rambam Health Care Campus approved this study protocol (0089-21-RMB), registered at clinicaltrials.gov (NCT05001594). The procedures are in accord with the ethical standards of the Helsinki Declaration. All of the participants gave their written informed consent to enter the study.

A single researcher, a physiotherapist with eight years of clinical experience and three years of experience in a motion analysis lab, conducted all measurements.

### Participants

The inclusion criteria comprised males and females aged 18-45 who had undergone ACLR surgery at Rambam Health Care Campus two months prior. Exclusion criteria included failure to provide informed consent, a history of previous ACL injury, prior lower limb injuries, and any active cardiovascular, neurological, or respiratory conditions.

Additionally, a control group comprising healthy participants was recruited. The inclusion criteria for the control group were healthy males and females aged 18-45 without any lower limb pain. The exclusion criteria were a previous lower limb surgery or injury, active cardiovascular, neurological, or respiratory conditions, and failure to provide informed consent.

### Research protocol

Using an online random number sequence generator, the participants were randomized into two groups; one group received an IVS device (*KneeMo^®^*), while the other group received a 3D-printed sham device without vibration. Both devices were positioned on the participant’s injured leg using straps above and below the knee, providing pressure. However, only the IVS device delivered external vibration stimulation. The vibration was synchronized to the gait cycle and activated just before the heel strike and is described in detail elsewhere.^14^ Due to the nature of the intervention, participants were not blinded, but the sham device was made to look and feel like the IVS device.

The participants first walked, ascended, and descended stairs without any device (baseline) and then walked and ambulated stairs with a device (IVS or sham). Each participant completed three repetitions of walking along a 20-meter corridor at three paces: self-selected normal, slow, and fast. The instructions for each condition were as follows and similar for all the participants. For the self-selected pace: "Walk across the corridor at your normal speed," for the slow pace: "Walk across the corridor at a slow speed," and for the fast pace: "Walk as fast as possible across the corridor".^15^ Participants then ascended and descended a 20-step staircase (rise= 17cm, run= 30cm) at their comfortable pace for three repetitions. The cycles of each participant were then averaged and used for the analysis.

Data were recorded at 120 Hz using seven IMU sensors (XSENS Awinda, Full citation.) attached to the participants’ lower limbs using Velcro straps: Upper leg (x2), lower leg (x2), feet (x2), and pelvis (x1).^16^ The Participants acclimated to the sensors before data collection, initially walking without any device (baseline phase) and then with either the IVS or sham device, depending on randomization (intervention phase).

The main outcomes included knee sagittal kinematics, with secondary outcomes focusing on hip and ankle sagittal kinematics (Figure 1).

**Figure 1.**
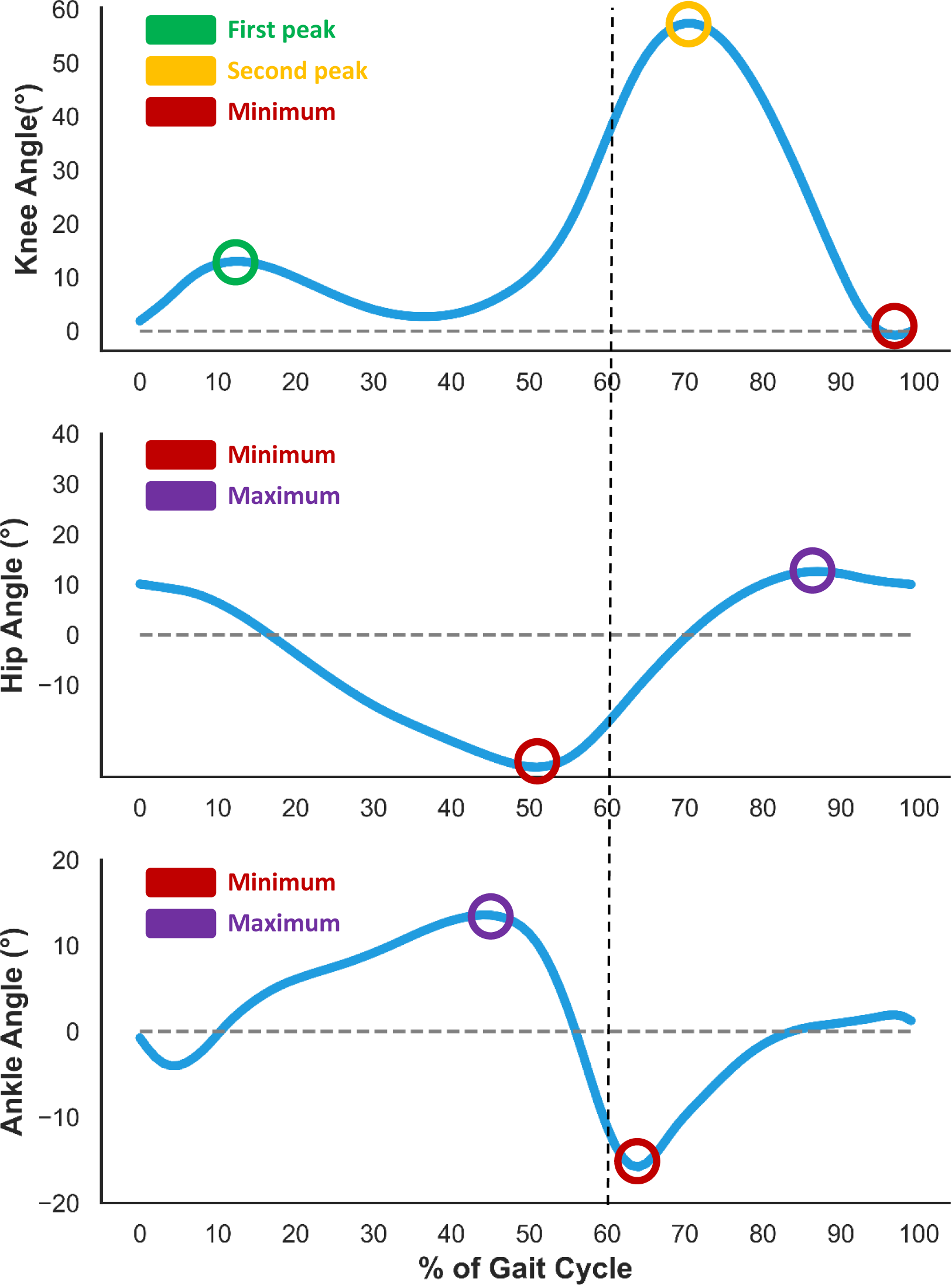
Points of interest during the gait cycle in the knee, hip and ankle joints. Gait cycle graphs for each of the lower limb joints with the points of interest marked for each of the joints.

### Data and statistical analysis

The normality of the data distribution was assessed using the Shapiro-Wilk test. Demographic differences between groups and between the injured and the left leg of the healthy controls were compared using the student t-test. Two-way mixed ANOVA tests evaluated between-group (group = IVS vs. sham device) and within-group (time = walking without a device - baseline phase. vs. walking with the IVS or sham device - intervention phase) differences of the different kinematic data, with significant interactions further evaluated using the ANOVA pairwise comparison. Effect sizes were reported using Partial Eta squared (partial η^2^).

An additional comparison was made between the ACLR cohort’s injured leg and the healthy participants’ left leg using an independent-sample t-test. The statistical significance was set at p<.05, and analyses were performed using IBM SPSS Statistics (Version 29).

A priori power analysis was conducted to determine the minimum sample size required to detect a medium effect based on an f-test and a within- and between-group interaction statistical test using G*Power (Version 3.1.9.7). The results indicated that the required sample size to achieve 80% power for detecting a medium effect (0.25), at a significance of α = .05, was n=34.

Due to the COVID-19 pandemic, data collection was slower and stopped earlier than planned. Therefore, the final sample size is smaller than the planned number. With 27 participants instead of 34, our planned type 2 error rate increased from 20% (80% power) to 30% (70% power).

The funding sources were not involved in the design, collection, analysis, and interpretation of data or the writing of this report.

## Results

### Participant characteristics

A total of 27 participants post-ACLR and 24 healthy controls were recruited between February 2022 and October 2023 (Table 1). The mean age of the ACLR participants was 23.5 ± 5.7 years, with a mean weight and height of 75.8 ± 11.6 kg and 177 ± 0.1 cm, respectively. Data for stair ambulation from one ACLR participant was corrupted, resulting in 26 participants being included in the stair ambulation analysis (Figure 2). The sagittal kinematics during gait and stair ascent of the injured and contralateral knee together with the left leg of the healthy participants are presented in Tables 2 and 3 and Appendices 1 and 2 for the hip and ankle.

**Figure 2.**
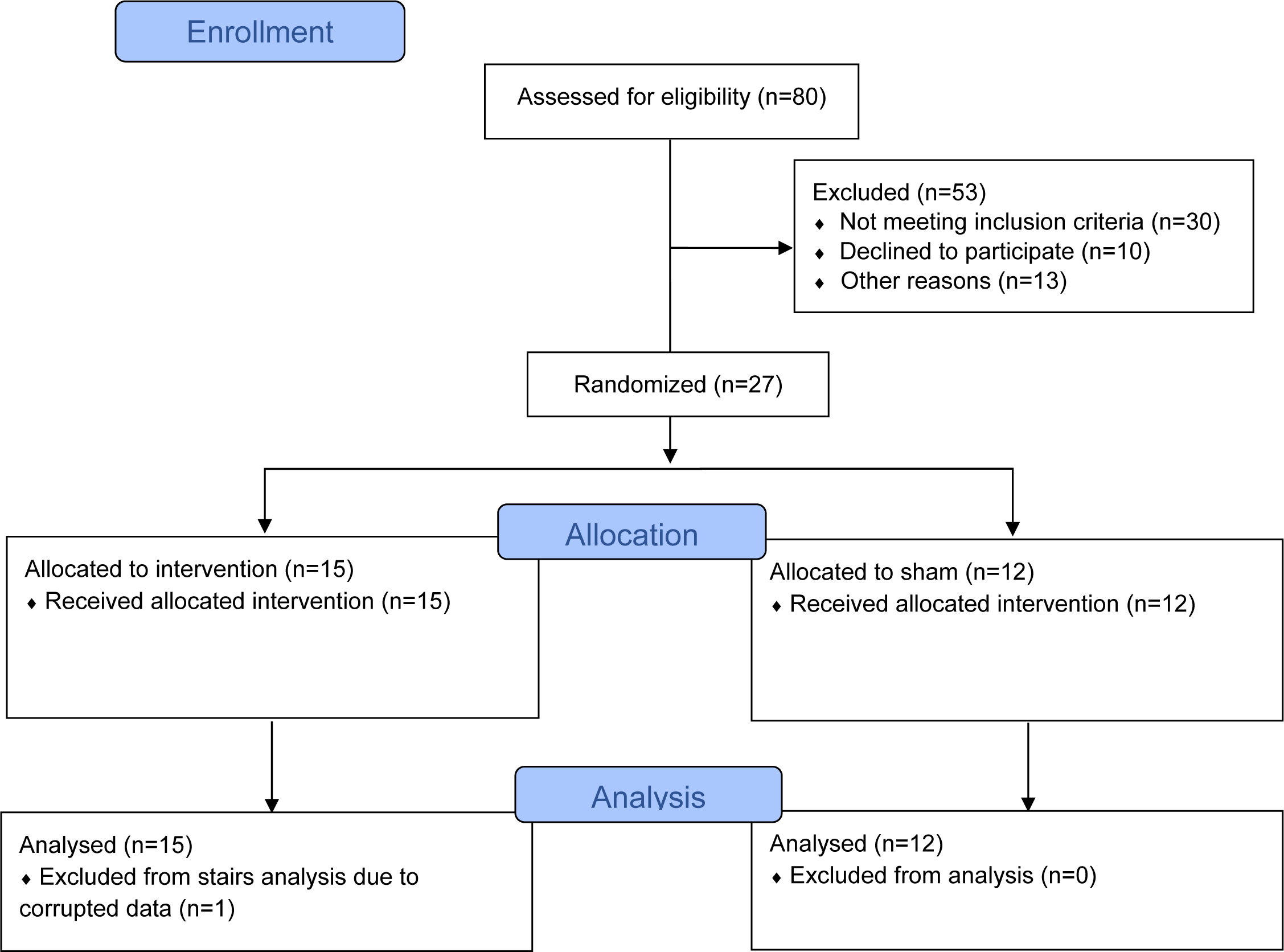
CONSORT Flow Diagram

**Table 1.**
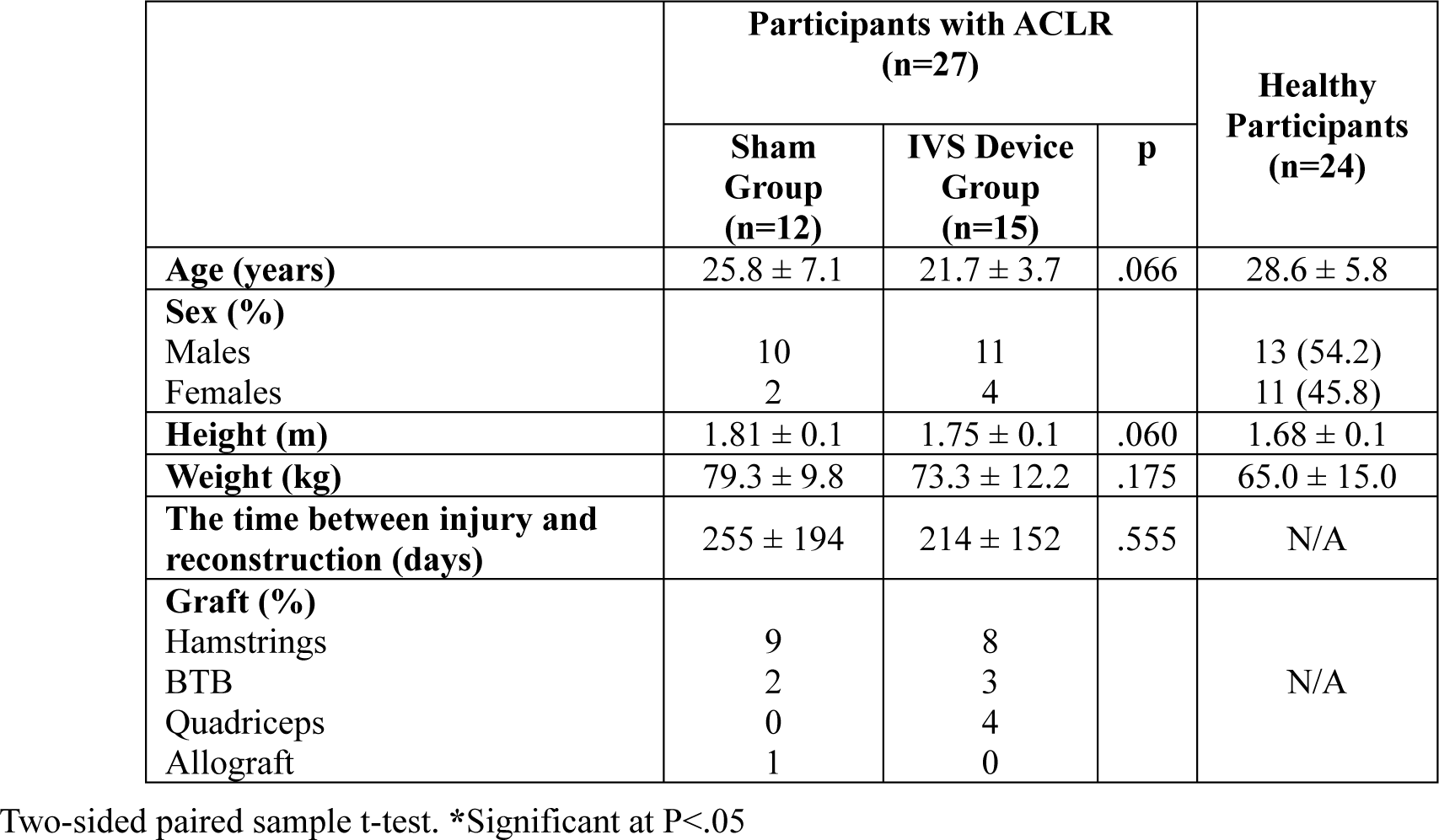
Demographics of the participants.

**Table 2.**
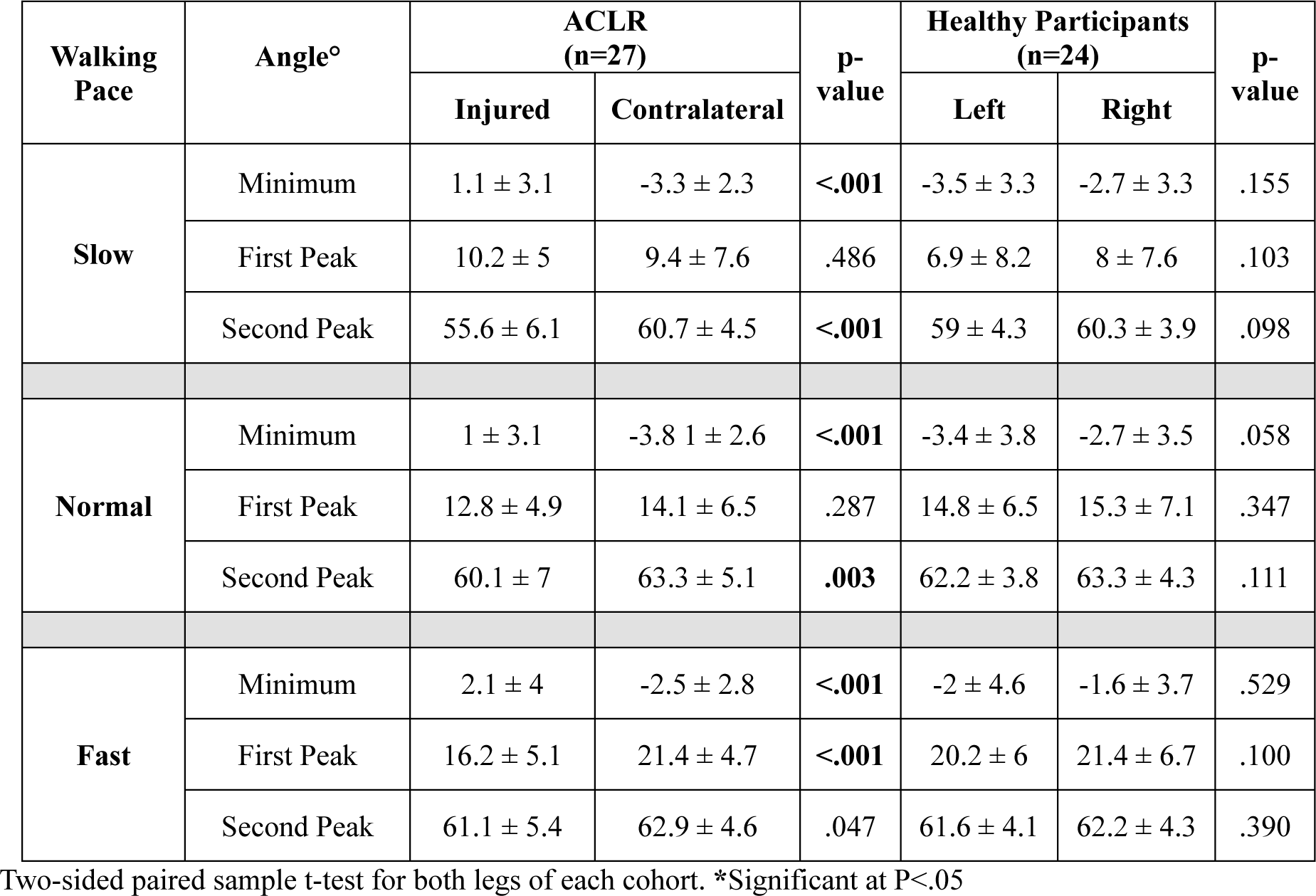
Knee sagittal angle of the participants while walking by group.

**Table 3.**
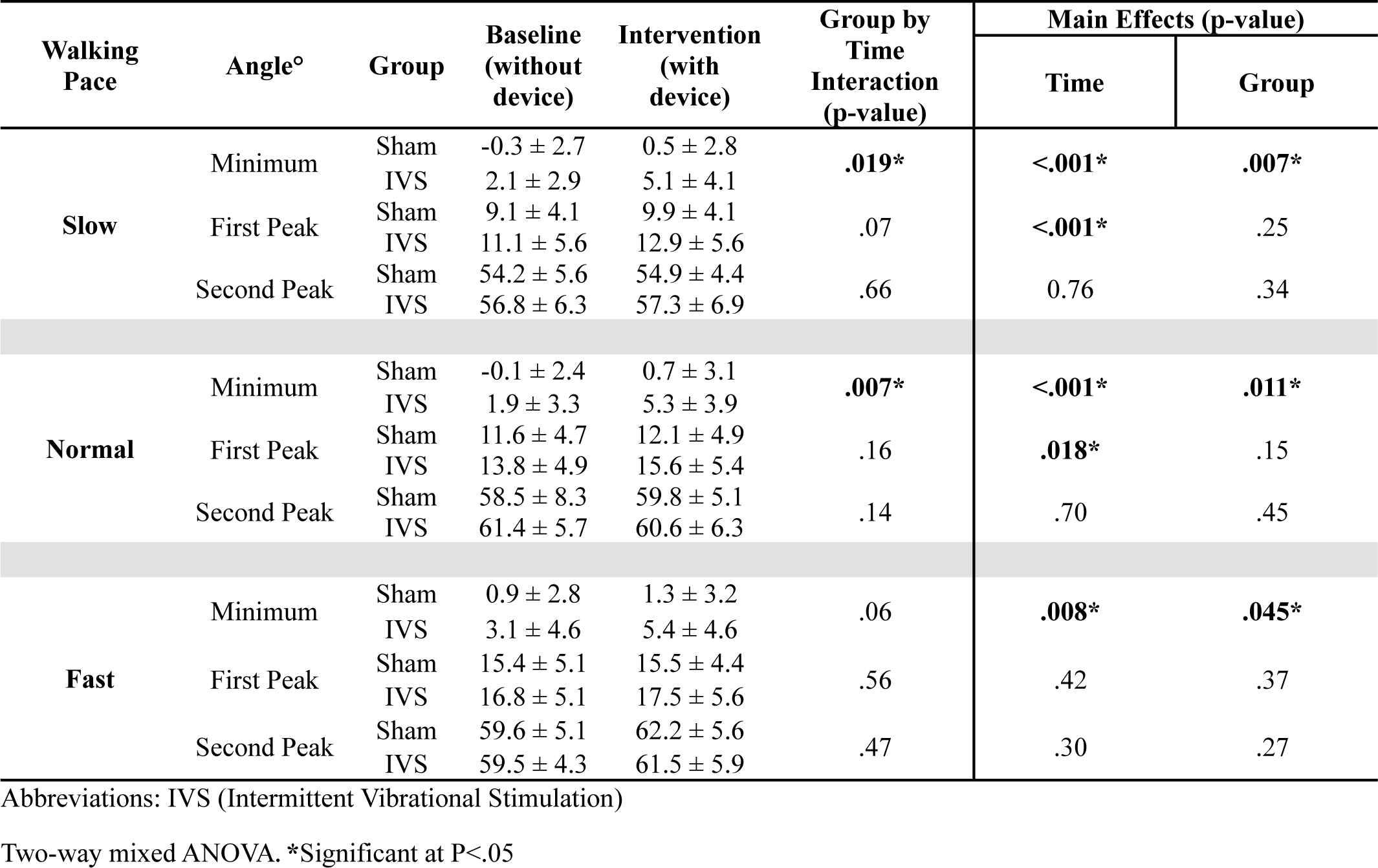
Baseline and Intervention Sessions, Angle Differences, and Effect Sizes for Knee Angles During Walking at Three Different Paces.

| Walking Pace | Angle° | Group | Baseline (without device) | Intervention (with device) | Group by Time Interaction (p-value) | Main Effects (p-value) |  |
| --- | --- | --- | --- | --- | --- | --- | --- |
|  |  |  |  |  |  | Time | Group |
| Slow | Minimum | Sham | -0.3 ± 2.7 | 0.5 ± 2.8 | **.019\*** | **<.001\*** | **.007\*** |
|  |  | IVS | 2.1 ± 2.9 | 5.1 ± 4.1 |  |  |  |
|  | First Peak | Sham | 9.1 ± 4.1 | 9.9 ± 4.1 | .07 | **<.001\*** | .25 |
|  |  | IVS | 11.1 ± 5.6 | 12.9 ± 5.6 |  |  |  |
|  | Second Peak | Sham | 54.2 ± 5.6 | 54.9 ± 4.4 | .66 | 0.76 | .34 |
|  |  | IVS | 56.8 ± 6.3 | 57.3 ± 6.9 |  |  |  |
| Normal | Minimum | Sham | -0.1 ± 2.4 | 0.7 ± 3.1 | **.007\*** | **<.001\*** | **.011\*** |
|  |  | IVS | 1.9 ± 3.3 | 5.3 ± 3.9 |  |  |  |
|  | First Peak | Sham | 11.6 ± 4.7 | 12.1 ± 4.9 | .16 | **.018\*** | .15 |
|  |  | IVS | 13.8 ± 4.9 | 15.6 ± 5.4 |  |  |  |
|  | Second Peak | Sham | 58.5 ± 8.3 | 59.8 ± 5.1 | .14 | .70 | .45 |
|  |  | IVS | 61.4 ± 5.7 | 60.6 ± 6.3 |  |  |  |
| Fast | Minimum | Sham | 0.9 ± 2.8 | 1.3 ± 3.2 | .06 | **.008\*** | **.045\*** |
|  |  | IVS | 3.1 ± 4.6 | 5.4 ± 4.6 |  |  |  |
|  | First Peak | Sham | 15.4 ± 5.1 | 15.5 ± 4.4 | .56 | .42 | .37 |
|  |  | IVS | 16.8 ± 5.1 | 17.5 ± 5.6 |  |  |  |
|  | Second Peak | Sham | 59.6 ± 5.1 | 62.2 ± 5.6 | .47 | .30 | .27 |
|  |  | IVS | 59.5 ± 4.3 | 61.5 ± 5.9 |  |  |  |

Abbreviations: IVS (Intermittent Vibrational Stimulation)
Two-way mixed ANOVA. \*Significant at P<.05

### Effects of vibration on the knee kinematics

#### Minimum knee flexion angle

As detailed in Table 3, statistically significant interactions between intervention and time were observed for knee minimum flexion angles during normal walking (F(1, 25) = 7.60, p = .007, partial η2 = .233) and slow walking (F(1, 25) = 6.32, p = .01, partial η2 = .208).

Post-hoc analysis revealed a statistically significant simple effect of group on the knee minimum flexion angle that was higher in the IVS group compared to the sham device at normal walking (mean difference = 4.60°, S.E. = 1.37°, p = .003, partial η^2^ =.309) and at slow walking (mean difference = 4.67°, S.E. = 1.40°, p = .003, partial η^2^ = .316). Moreover, there was a statistically significant difference at fast walking with a higher knee minimum flexion angle in the IVS group (mean difference = 4.04°, S.E. = 1.57°, p = .017, partial η^2^ = .208).

Furthermore, there was a statistically significant simple effect of time on the knee minimum flexion angle that was higher during the intervention phase in the IVS group at normal walking (mean difference = 3.44°, S.E. = 0.73°, p < .001, partial η^2^ =.610) and at slow walking (mean difference = 3.14°, S.E. = 0.72°, p < .001, partial η^2^ =.588). Similarly, a significant difference was found in fast walking during the intervention phase in the IVS group (mean difference = 2.25°, S.E. = 0.81°, p = .015, partial η^2^ =.354).

An independent-sample t-test was conducted to assess whether there were differences in knee minimum flexion angles between the ACLR IVS group’s injured leg and the healthy participants’ left leg. The results revealed that during the baseline phase, while walking at a normal pace, the injured knee of the ACLR group exhibited greater flexion (1.9° ± 3.4) compared to the knee of healthy participants (-3.4° ± 3.8), a statistically significant mean difference of 5.3° (95% CI [2.9, 7.7], p < .001). Similar differences were observed at both slow walking (mean difference of 5.6°, 95% CI [3.5, 7.7], p < .001) and fast walking (mean difference of 5.1°, 95% CI [2.0, 8.2], p = .002).

During the intervention phase, the injured knee of the ACLR group was more flexed (5.3° ± 3.8) compared to the healthy participants, with a statistically significant mean difference of 8.8° (95% CI [6.2, 11.3], p < .001). The same was true at both slow walking (mean difference of 8.6°, 95% CI [6.2, 11.1], p < .001) and at fast walking (mean difference of 7.4°, 95% CI [4.3, 10.4], p = .002).

### First peak knee flexion angle

No statistically significant interaction was found between intervention and time for the first peak flexion angles at any walking speed. However, there was a statistically significant main effect of time that showed a difference in the first peak flexion angle at normal walking speed (F(1, 25) = 6.47, p = .018, partial η2 = .206) and at slow walking speed (F(1, 25) = 14.59, p < .001, partial η2 = .378).

Post-hoc analysis revealed a statistically significant simple effect of time on the first peak flexion angle that was higher at the intervention phase in the IVS group at normal walking speed (mean difference = 1.77°, S.E. = 0.76°, p = .036, partial η^2^ =.278). A statistically significant simple effect of time was also found at slow walking speed both in the sham device group (mean difference = 0.80°, S.E. = 0.29°, p = .021, partial η^2^ =.396) and in the IVS group (mean difference = 2.35°, S.E. = 0.72°, p = .006, partial η^2^ =.452).

An independent-sample t-test found no significant differences between the injured leg of the ACLR IVS group participants and the healthy participants’ left leg.

### Second peak knee flexion angle

There was no statistically significant interaction between the intervention and time on second peak flexion angles across all walking speeds. Furthermore, no statistically significant main effects of time or group were observed. An independent-sample t-test revealed no significant differences between the injured leg of the ACLR IVS device group and the left leg of the healthy participants.

### Stairs ambulation

Table 4 shows a statistically significant interaction between the intervention and time concerning the knee maximum angle (F(1, 25) = 5.88, p = .023, partial η2 = .197). Post-hoc tests revealed a statistically significant simple effect of time on the knee maximum angle, indicating a decrease during the intervention phase in the IVS group (mean difference = 1.88°, S.E. = 0.69°, p = .018, partial η2 =.362).

**Table 4.** Baseline and Intervention Sessions, Angle Differences, and Effect Sizes for the Lower Limbs Angles During Stair Ascent.

| Joint | Angle° | Group | Baseline<br>(without<br>device) | Intervention<br>(with<br>device) | Group by<br>Time<br>Interaction<br>(p-value) | Main Effects (p-value) |  |
| --- | --- | --- | --- | --- | --- | --- | --- |
|  |  |  |  |  |  | Time | Group |
| Hip | Minimum | Sham | 13.7 ± 6.9 | 14 ± 8.4 | .47 | .73 | .52 |
|  |  | IVS | 11.9 ± 11.2 | 11.1 ± 10 |  |  |  |
|  | Maximum | Sham | 64.8 ± 7.7 | 65 ± 8.5 | .23 | .33 | .45 |
|  |  | IVS | 63.2 ± 9.1 | 61.5 ± 9.5 |  |  |  |
| Knee | Minimum | Sham | 13.9 ± 4.9 | 14.5 ± 4.8 | .18 | .019* | .85 |
|  |  | IVS | 12.9 ± 4.5 | 14.9 ± 5.1 |  |  |  |
|  | Maximum | Sham | 83.2 ± 7.8 | 83.5 ± 7.9 | .023* | .093 | .47 |
|  |  | IVS | 82.2 ± 6.8 | 80.3 ± 6.7 |  |  |  |
| Ankle | Minimum | Sham | -6.8 ± 5.8 | -7.4 ± 7.4 | .037* | .10 | .55 |
|  |  | IVS | -11.2 ± 6.9 | -6.6 ± 10.4 |  |  |  |
|  | Maximum | Sham | 22.7 ± 3.9 | 23.1 ± 6.3 | .13 | .041* | .91 |
|  |  | IVS | 21.3 ± 4.5 | 24.1 ± 5.7 |  |  |  |

Abbreviations: IVS (Intermittent Vibrational Stimulation)
Two-way mixed ANOVA. \*Significant at P<.05

Furthermore, a significant main effect of time emerged regarding the knee minimum angle (F(1, 25) = 6.31, p = .019, partial η2 = .208). Post-hoc tests revealed a significant simple effect of time on the knee minimum angle (mean difference = 2.06°, S.E. = 0.93°, p = .047, partial η2 =.271) indicating an increase during the intervention phase in the IVS group.

There were no statistically significant interactions or main effects during stair descent.

### Effects of vibration on the ankle kinematics

There were no statistically significant interactions for the ankle kinematics across any of the walking measurements (Table 5). However, a significant main effect of time was identified, showing differences in the ankle maximum angle at slow (F(1, 25) = 8.54, p = .007, partial η2 = .263) and fast walking paces (F(1, 25) = 11.14, p = .003, partial η2 = .308).

**Table 5.** Baseline and Intervention Sessions, Angle Differences, and Effect Sizes for Ankle Angles During Walking at Three Different Paces.

| Walking Pace | Angle° | Group | Baseline<br>(without<br>device) | Intervention<br>(with<br>device) | Group by Time<br>Interaction<br>(p-value) | Main Effects (p-value) |  |
| --- | --- | --- | --- | --- | --- | --- | --- |
|  |  |  |  |  |  | Time | Group |
| Slow | Minimum | Sham | -15.5 ± 4.8 | -14.8 ± 3.6 | .24 | .68 | .91 |
|  |  | IVS | -14.1 ± 7.9 | -15.6 ± 8.5 |  |  |  |
|  | Maximum | Sham | 16.6 ± 2.1 | 17 ± 2.1 | .37 | **.007\*** | .72 |
|  |  | IVS | 16.1 ± 2.4 | 16.9 ± 2.3 |  |  |  |
| Normal | Minimum | Sham | -19.4 ± 6.5 | -19.7 ± 6.1 | .74 | .51 | .78 |
|  |  | IVS | -19.7 ± 8.6 | -20.7 ± 5.9 |  |  |  |
|  | Maximum | Sham | 16.3 ± 2.4 | 15.7 ± 3.2 | .29 | .13 | .71 |
|  |  | IVS | 16.4 ± 2.7 | 16.3 ± 2.8 |  |  |  |
| Fast | Minimum | Sham | -23.1 ± 7.6 | -22.5 ± 8.7 | .33 | .98 | .44 |
|  |  | IVS | -24.5 ± 5.7 | -25 ± 4.9 |  |  |  |
|  | Maximum | Sham | 14.7 ± 2.9 | 15 ± 3.1 | .09 | **.003\*** | .57 |
|  |  | IVS | 13.7 ± 3.4 | 14.6 ± 3.4 |  |  |  |

Abbreviations: IVS (Intermittent Vibrational Stimulation)
Two-way mixed ANOVA. \*Significant at P<.05

Post-hoc analyses revealed a significant simple effect of time on the ankle maximum angle for slow walking (mean difference = 0.77°, S.E. = 0.30°, p = .023, partial η2 =.338) and fast walking (mean difference = 0.93°, S.E. = 0.30°, p = .009, partial η2 =.396), with higher angles observed during the intervention phase in the IVS group.

During the baseline phase, while walking at a slow pace, the ankles of the ACLR group’s injured legs exhibited less plantarflexion (-14.0° ± 7.9) compared to those of healthy participants (-19.1° ± 7.3), resulting in a statistically significant mean difference of 5.1° (95% CI [0.1, 10.1], p = .04). Conversely, during the intervention phase, the ankles of the ACLR group’s injured legs demonstrated more dorsiflexion (16.8° ± 2.3) compared to healthy participants (14.6° ± 3.1), with a statistically significant mean difference of 2.2° (95% CI [0.2, 4.2], p = .02).

### Stair ambulation

As seen in Table X, there was a statistically significant interaction between the intervention and time on the ankle minimum angle (F(1, 25) = 4.86, p = .037, partial η2 = .168). Post-hoc tests revealed a statistically significant simple effect of time on the ankle minimum angle (mean difference = 4.58°, S.E. = 1.91°, p = .032, partial η2 =.306), indicating a higher angle during the intervention phase in the IVS group.

Furthermore, there was a statistically significant main effect of time regarding the ankle maximum angle (F(1, 25) = 4.65, p = .041, partial η2 = .162). Post-hoc tests found a statistically significant simple effect of time on the ankle maximum angle (mean difference = 2.86°, S.E. = 1.17°, p = .030, partial η2 =.313) that was higher at the intervention phase in the IVS group.

There were no statistically significant interactions or main effects during stair descent.

### Effects of vibration on the hip kinematics

There were no statistically significant interactions or main effects for the hip kinematics in any measurements (Appendix 3 and 4) during walking or stair ambulation. An independent-sample t-test found no significant differences between the injured leg of the ACLR IVS group and the healthy participants’ left leg.

### Effects of IVS on the lower limb kinematics

As detailed in Table 6, IVS is associated with changes in lower limb kinematics across various activities. For knee kinematics, the minimum knee flexion angles during slow, normal, and fast walking paces increased by 4.67°, 4.60°, and 4.04°, respectively. During stair ascent, the minimum knee flexion angle also increased by 2.06°, whereas the peak knee flexion angle decreased by 1.88°.

**Table 6.** The Effects of intermittent muscle vibration on the lower limb kinematics during walking at different paces and stair ambulation.

| Variables<br>(Angle°) | Walking |  |  | Stair ambulation |  |
| --- | --- | --- | --- | --- | --- |
|  | Slow | Normal | Fast | Ascent | Descent |
| Knee |  |  |  |  |  |
| Minimum | ↑ 4.67° | ↑ 4.60° | ↑ 4.04° | ↑ 2.06° | — |
| First peak | ↑ 2.35° | ↑ 1.77° | — | ↓ 1.88° | — |
| Second peak | — | — | — | N/A |  |
| Hip |  |  |  |  |  |
| Minimum | — | — | — | — | — |
| Maximum | — | — | — | — | — |
| Ankle |  |  |  |  |  |
| Minimum | — | — | — | ↑ 4.58° | — |
| Maximum | ↑ 0.77° | — | ↑ 0.93° | ↑ 2.86° | — |

↑ = increased angle. ↓ = decreased angle.

Similarly, the ankle joint increased the maximum angle during slow and fast walking paces (0.77° and 0.93°) and both the minimum and maximum angles during stair ascent (4.58° and 2.86°). On the contrary, the hip kinematics did not show significant changes in any of the evaluated activities.

## Discussion

We investigated the effects of IVS on lower limb kinematics in individuals two months post-ACLR. Our findings demonstrated that IVS increased the knee’s minimum flexion angle across slow, normal, and fast walking paces. Further, IVS significantly increased the first peak knee flexion angle during the slow and normal paces.

Given that the typical duration of the gait cycle is approximately 0.98-1.07 seconds,^17^ the observed difference between the minimum knee flexion angle just before heel strike and the first peak at weight acceptance is approximately 100 milliseconds. We speculate that IVS might cause a stronger quadriceps contraction and has a pain reduction effect during the stance phase, where the participant has his weight on the injured leg, lasting from heel strike to weight acceptance.^18^ Therefore, future investigations should assess the effects of different timings of the IVS to induce vibration at a specific location during the gait cycle.

Comparing this study’s findings with those of other studies, our results align with a previous study that utilized IVS for knee pathologies^9^, albeit in an older population with mixed knee pathologies. This study reported a trend toward higher knee flexion during the loading response (16.4°±5.5 control, 16.9°±5.3 IVS), although it did not specifically assess the minimum knee flexion angle. The bigger differences in the current study may be due to the more acute nature of ACLR.

Further, another study examining different vibration modalities’ effects on muscle activation during squatting demonstrated improved quadriceps activation among patients four years post-ACLR, highlighting the applicability in this population. However, this study didn’t report on the kinematic changes of the lower limb following the IVS.^19^. In alignment with our research, Blackburn et al. (2020) demonstrated that vibration therapy could effectively improve gait biomechanics linked to posttraumatic knee osteoarthritis, further supporting the idea of the potential of different vibrational interventions in enhancing long-term joint health post-ACL reconstruction.^20^

Next, we found kinematic changes during stair ascent while using the IVS device, with a significantly higher minimum flexion angle compared to the sham group. This finding contrasts a previous study by Fischer et al. (2021), who observed no changes in the knee flexion angle during stair ascent and a reduction in the knee flexion angle during stair descent among patients with knee pain.^14^ This inconsistency may be due to their study’s older and heterogenous population.

The clinical implications of the observed changes in knee kinematics are significant for individuals undergoing post-ACLR rehabilitation, suggesting a potential for IVS to improve quadriceps avoidance gait, potentially reducing the risk of posttraumatic osteoarthritis by enhancing the knee’s ability to absorb force during weight acceptance.

### Study Limitations

It is important to acknowledge this study’s limitations. It is important to note that the findings are from a relatively small sample size, which may affect the generalizability of the results to a broader population. However, the statistical significance achieved despite the small cohort underscores the potential efficacy of the intervention. Next, the lack of blindness may introduce bias. In our study, the nature of the intervention—where participants were aware of the device’s activation (IVS or sham)—presents a potential limitation. Still, our analysis focused on objective kinematic measurements, which are less susceptible to bias.

Further, this study primarily evaluated the immediate kinematic effects of IVS, providing insights into early rehabilitation outcomes. However, the long-term clinical significance of these changes remains unexplored. Additionally, while inertial measurement sensors offer ecological validity, they lack optimal validity compared to camera-based motion capture systems; therefore, healthy controls are included for accurate comparisons.^16^

Future research with larger sample sizes is essential to validate these findings, explore the intervention’s applicability across diverse populations and activities, and focus on the long-term effects and clinical implications of IVS in ACLR populations.

### Conclusions

IVS around the knee and synced to the gait cycle increase knee flexion during gait at different speeds and stair ascent. Further research is needed to explore the long-term effects of IVS and the clinical meaning of these changes.

## Data Availability

All data produced in the present study are available upon reasonable request to the authors

## Abbreviations

ACL: (Anterior cruciate ligament)
ACLR: (Anterior cruciate ligament reconstruction)
IMU: (Inertial measurement unit)
IVS: (Intermittent vibrational stimulation )

## Acknowledgments

KneeMo devices were provided by SDI SomaTX Design Inc.

## Suppliers

SDI SomaTX Design Inc. Minden, Nevada

---

**Appendix 1.** Hip sagittal angle of the participants by group.

| Walking Pace | Angle° | ACLR (n=27) |  | p value | Healthy Participants (n=24) |  | p-value |
| --- | --- | --- | --- | --- | --- | --- | --- |
|  |  | Injured | Contralateral |  | Left | Right |  |
| Slow | Minimum | -9.2 ± 5.3 | -11.3 ± 5.4 | **.031\*** | -10.5 ± 8.4 | -11.6 ± 5.3 | .578 |
|  | Maximum | 24 ± 4.6 | 26.2 ± 4.4 | **<.001\*** | 24.7 ± 5.3 | 25.9 ± 4.5 | **.017\*** |
| Normal | Minimum | -11.1 ± 5.8 | -13.5 ± 5.6 | **.017\*** | -13.8 ± 4.8 | -13.3 ± 4.7 | .237 |
|  | Maximum | 27 ± 4.9 | 28.7 ± 4.3 | **.001\*** | 29 ± 5.5 | 29.5 ± 4.8 | .340 |
| Fast | Minimum | -11.8 ± 6.2 | -15.1 ± 6.1 | **.003\*** | -15.4 ± 5.1 | -14.9 ± 4.5 | .392 |
|  | Maximum | 31 ± 6.1 | 32.6 ± 5.5 | **.008\*** | 34.4 ± 7 | 34.8 ± 6.6 | .297 |

Two-sided paired sample t-test for the right and left leg of each cohort.
Two-way mixed ANOVA. \*Significant at P<.05

**Appendix 2.**
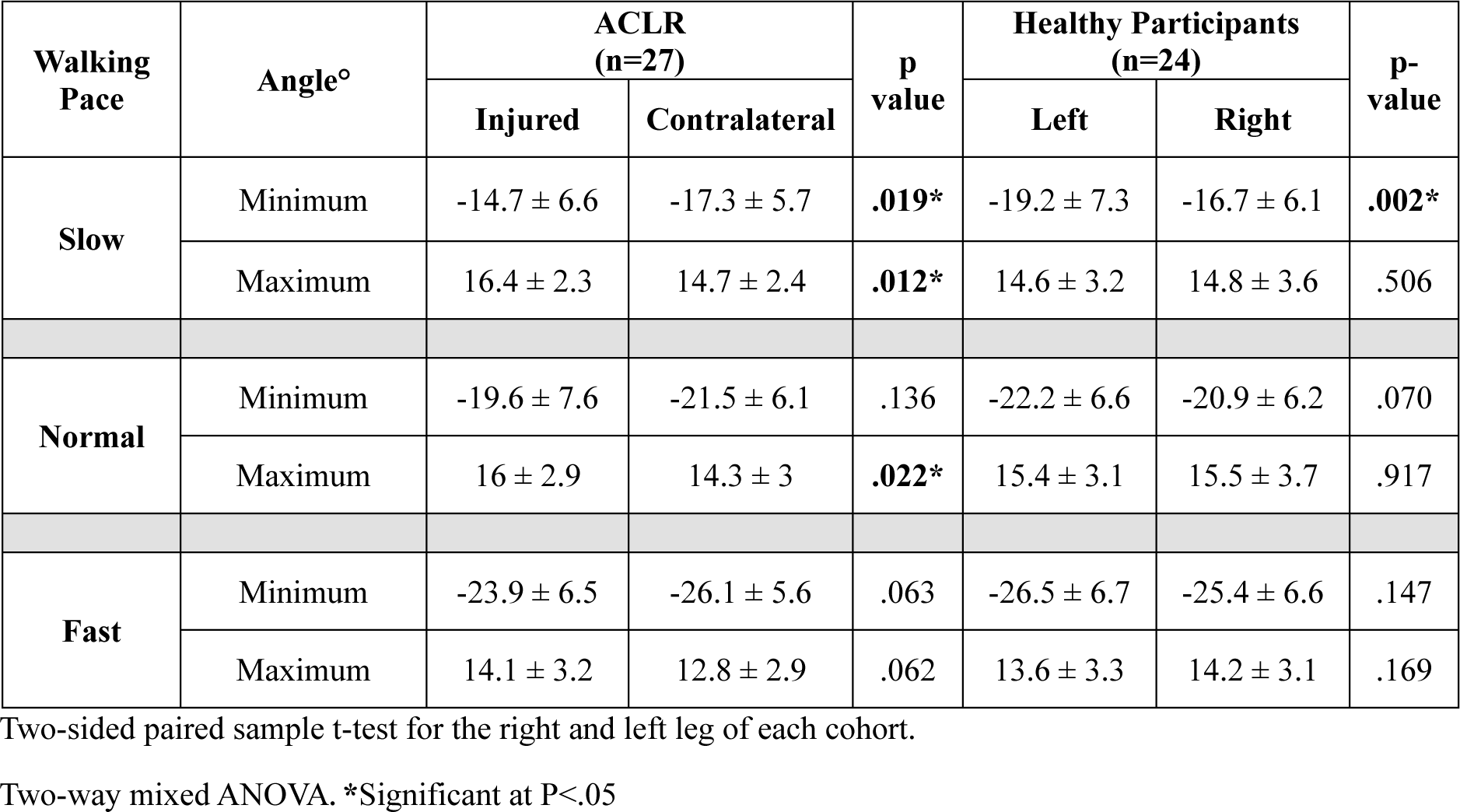
Ankle sagittal angle of the participants by group.

| Walking Pace | Angle° | ACLR (n=27) |  | p value | Healthy Participants (n=24) |  | p-value |
| --- | --- | --- | --- | --- | --- | --- | --- |
|  |  | Injured | Contralateral |  | Left | Right |  |
| Slow | Minimum | -14.7 ± 6.6 | -17.3 ± 5.7 | .019* | -19.2 ± 7.3 | -16.7 ± 6.1 | .002* |
|  | Maximum | 16.4 ± 2.3 | 14.7 ± 2.4 | .012* | 14.6 ± 3.2 | 14.8 ± 3.6 | .506 |
| Normal | Minimum | -19.6 ± 7.6 | -21.5 ± 6.1 | .136 | -22.2 ± 6.6 | -20.9 ± 6.2 | .070 |
|  | Maximum | 16 ± 2.9 | 14.3 ± 3 | .022* | 15.4 ± 3.1 | 15.5 ± 3.7 | .917 |
| Fast | Minimum | -23.9 ± 6.5 | -26.1 ± 5.6 | .063 | -26.5 ± 6.7 | -25.4 ± 6.6 | .147 |
|  | Maximum | 14.1 ± 3.2 | 12.8 ± 2.9 | .062 | 13.6 ± 3.3 | 14.2 ± 3.1 | .169 |

Two-sided paired sample t-test for the right and left leg of each cohort.
Two-way mixed ANOVA. \*Significant at P<.05

**Appendix 3.** Baseline and Intervention Sessions, Angle Differences, and Effect Sizes for Hip Angles During Walking at Three Different Paces.

| Walking Pace | Angle° | Group | Baseline<br>(without<br>device) | Intervention<br>(with<br>device) | Group by Time<br>Interaction<br>(p-value) | Main Effects (p-value) |  |
| --- | --- | --- | --- | --- | --- | --- | --- |
|  |  |  |  |  |  | Time | Group |
| Slow | Minimum | Sham | -9.4 ± 5.2 | -9.8 ± 5.1 | .55 | .55 | .91 |
|  |  | IVS | -9.1 ± 5.6 | -10.3 ± 5.8 |  |  |  |
|  | Maximum | Sham | 24.4 ± 4.6 | 24.7 ± 4.7 | .93 | .42 | .48 |
|  |  | IVS | 23.5 ± 4.7 | 23.5 ± 5.1 |  |  |  |
| Normal | Minimum | Sham | -10.9 ± 5.7 | -11.7 ± 5.2 | .42 | .47 | .96 |
|  |  | IVS | -11.2 ± 6.1 | -11.2 ± 5.6 |  |  |  |
|  | Maximum | Sham | 27.9 ± 4.5 | 27.6 ± 4.5 | .39 | .77 | .51 |
|  |  | IVS | 26.2 ± 5.2 | 26.7 ± 6.1 |  |  |  |
| Fast | Minimum | Sham | -11.3 ± 5.7 | -12.1 ± 5.4 | .57 | .27 | .81 |
|  |  | IVS | -12.2 ± 6.7 | -12.4 ± 6.4 |  |  |  |
|  | Maximum | Sham | 31.8 ± 5.4 | 31.1 ± 5.7 | .68 | .31 | .63 |
|  |  | IVS | 30.4 ± 6.7 | 30.1 ± 7.1 |  |  |  |

Abbreviations: IVS (Intermittent Vibrational Stimulation)
Two-way mixed ANOVA. \*Significant at P<.05

**Appendix 4.** Baseline and Intervention Sessions, Angle Differences, and Effect Sizes for the Lower Limbs Angles During Stair Descent (n=26)

| Joint | Angle° | Group | Baseline<br>(without<br>device) | Intervention<br>(with<br>device) | Group by Time<br>Interaction<br>(p-value) | Main Effects (p-value) |  |
| --- | --- | --- | --- | --- | --- | --- | --- |
|  |  |  |  |  |  | Time | Group |
| Hip | Minimum | Sham | 13.1 ± 5.1 | 13.4 ± 5.1 | .22 | .45 | .75 |
|  |  | IVS | 12.8 ± 10.9 | 11.7 ± 10.4 |  |  |  |
|  | Maximum | Sham | 34.7 ± 7.1 | 36.2 ± 6.4 | .06 | .96 | .35 |
|  |  | IVS | 33.1 ± 9.5 | 31.6 ± 10.2 |  |  |  |
| Knee | Minimum | Sham | 11.6 ± 4.9 | 12.8 ± 4.8 | .64 | .10 | .85 |
|  |  | IVS | 12.3 ± 5.3 | 12.9 ± 5.4 |  |  |  |
|  | Maximum | Sham | 78.7 ± 14.4 | 82.9 ± 7.8 | .10 | .38 | .88 |
|  |  | IVS | 80.9 ± 8.2 | 79.6 ± 10.2 |  |  |  |
| Ankle | Minimum | Sham | -25 ± 5.3 | -24.8 ± 5.2 | .87 | .69 | .46 |
|  |  | IVS | -23.4 ± 5.7 | -23.1 ± 6.4 |  |  |  |
|  | Maximum | Sham | 27.1 ± 7.6 | 28.3 ± 5.5 | .27 | .84 | .95 |
|  |  | IVS | 28.3 ± 5.5 | 27.4 ± 8.6 |  |  |  |

Abbreviations: IVS (Intermittent Vibrational Stimulation)
Two-way mixed ANOVA. \*Significant at P<.05

## Notes

**Funding statement:** Arielle Fischer was supported by the Zuckerman STEM Leadership Program. In addition, this research was supported by the Israel Science Foundation (grant No. 2070658).

### Competing Interest Statement

Arielle Fischer is on the SDI advisory team.

### Clinical Trial

NCT05001594

### Funding Statement

Arielle Fischer was supported by the Zuckerman STEM Leadership Program. In addition, this research was supported by the Israel Science Foundation (grant No. 2070658).

### Author Declarations

Ethics committee of Rambam Health Care Campus gave ethical approval for this work (0089-21-RMB)

